# Predictors of Affective and Cognitive Alterations in Minor Stroke Patients: clinical, omics, neuropsychological and radiological signatures

**DOI:** 10.1101/2023.08.03.23293629

**Authors:** Cristina Pereira, Gerard Mauri, Daniel Vázquez-Justes, Raquel Mitjana, Gerard Piñol, Gloria Arqué, Francisco Purroy

**Affiliations:** Clinical Neurosciences research group, Institut de Recerca Biomèdica de Lleida, Lleida, Spain; Stroke Unit. Department of Neurology. Hospital Universitari Arnau de Vilanova de Lleida, Spain; Institute of Diagnostic Imaging (IDI), Hospital Universitari Arnau de Vilanova de Lleida, Spain; Cognitive Disordes Unit (UTC), Hospital Santa Maria de Lleida, Spain; Psychology Department, Universitat de Lleida (UdL), Lleida, Spain; Medicine Department, Universitat de Lleida (UdL), Lleida, Spain

**Keywords:** minor Stroke, affective alterations, cognitive alterations, post-stroke depression, observational study, neuropsychological battery, biomarker discovery, neuroimaging, metabolomics, lipidomics, prognosis

## Abstract

Background. Despite the mild severity of initial symptoms, minor stroke (MS) is a common clinical condition that can be associated with significant affective and cognitive alterations. Methods. The PSICOICTUS project aimed to investigate the predictors of these alterations in a cohort of 118 consecutive MS patient. This observational, longitudinal, and prospective study included comprehensive evaluation at baseline (within the first five days of symptom onset) and follow-ups at 15 days, 6 months, and 12 months. A screening battery consisting of the Montreal Cognitive Assessment (MoCA), Montgomery-Åsberg Depression Rating Scale (MADRS), and Apathy Evaluation Scale-Clinician version (AES-c) was used to identify patients with affective and/or cognitive alterations. Results. Cognitive alterations were further assessed using a comprehensive neuropsychological battery. Screening tests revelated that 17.0% of patients had affective alterations, 9.3% had cognitive alterations, and 14.4% presented with both affective and cognitive alterations. Among patients with cognitive alterations, executive functions, attention and processing speed were found to be the most affected domains. The study also concluded a biomarker discovery analysis involving MRI-based neuroimaging and untargeted metabolomics/lipidomics analysis. Several predictors of affective and cognitive alterations were identified. A history of previous depressions, lower levels of Isoleucyl-Isoleucine, and PC (38:4) were associated with affective alterations. On the other hand, age above 70, the presence of a middle cerebral artery ischemic lesion, higher creatinine levels, lower triglyceride levels, and higher concentrations of 2-hydroxyhexadecanoylcarnitine were predictive of cognitive alterations. Female sex, previous history of depressive syndrome, and a higher number of chronic strokes were identified as predictors for both affective and cognitive alterations. Conclusions. These findings highlight the importance of considering affective and cognitive alterations in the management of patients with MS. The identification of specific predictors would improve the development of a differential management approach tailored to the needs of MS patients.

## INTRODUCTION

Patients with minor stroke (MS) are characterized by presenting mild initial severity. Surely, for this reason are not included in the main clinical trials of reperfusion therapies. However, despite the clinical symptoms observed during the acute phase in MS patients, sequelae can occur in the chronic phase, which can interfere with daily activities and contribute to post-stroke disability. Depression in the most frequent affective complication after stroke [1] and a significant factor in predicting poor functional recovery [2]. Specifically, within the first year of follow-up, approximately 30% of MS patients developed depression [3]. Although apathy has been less evaluated, various research studies have found it to be a prevalent complication after stroke [4]. The prevalence of apathy in MS patients ranges from 19% and 50%, depending on the sample, assessment timing, and evaluation scale used [5]. Understanding the spectrum of complications following a MS could guide management decisions in the acute phase, promoting reperfusion treatments and during the follow-up.

Extensive research has been conducted on cognitive impairment following ischemic stroke, with prevalence rates ranging from 35.2% to 87% of patients [6]. Non-amnesic multiple domain impairment is the most closely associated with vascular dementia [7]. While magnetic resonance imaging (MRI) variables are crucial for diagnosing MS based on tissue definition [8], there is currently limited information available regarding neuroimaging data and the development of affective and cognitive alterations after MS [9]. On the other hand, in recent years, the use of metabolomics and lipidomics techniques has facilitated the search for plasma biomarkers associated with affective and/or cognitive screening after MS. This research is of significant interest in identifying new diagnostic and prognostic predictors. Existing studies have primarily focused on patients with ischemic stroke who scored high on the National Institute Health Stroke Scale (NIHSS) when describing risk predictors of these alterations.

Our objective was to recruit a cohort of MS patients to assess their clinical and neuropsychological profile, determine the radiological signature and define circulating biomarker of affective and cognitive alterations following MS.

## MATERIALS AND METHODS

### Participants and study design

A registry-based cohort study was conducted following the STROBE guidelines [10]. MS patients were prospectively recruited from January 2018 to March 2020 at the Hospital universitari Arnau de Vilanova (HUAV, Lleida, Spain). The local ethics committee approved the study (HUAV, code: 1838), and written informed consent was obtained from all participants.

MS was defined as an ischemic stroke with a score of ≤5 on the NIHSS and an evidence of new brain ischemia on MRI (tissue-based definition of stroke [8]). The study included patients aged 18 to 85 years old with a modified Ranking scale score (mRS) of ≤3. Patients with mild cognitive impairment (MCI) or dementia diagnosed before the stroke event, language barriers, illiteracy, severe language impairment defined by a score greater than 2 on the language item on the NIHSS scale, severe visual or auditory disabilities and severe medical comorbidity were excluded. All recruited patients were reviewed by a senior neurologist (FP).

### Procedures

At baseline time (3-5 days from the onset of symptoms), patients underwent a semi-structured interview to collect the clinical, socio-demographic, neurological, and pharmacological variables. Affective and cognitive screening batteries were administered at this time to assess depression, apathy, and cognitive impairment.

Depression was assessed using the Montgomery-Åsberg Depression Rating Scale (MADRS) [11] with a cut-off score of 7. Apathy was evaluated using the clinician version of the Apathy Evaluation Scale (AES-C) [12] with a cut-off score of 37 [13]. Cognitive impairment was assessed using the Montreal Cognitive Assessment (MoCA) [14] with a cut-off of 24, where higher scores indicate better outcomes. Patients who screened positive for cognitive impairment underwent a complete neuropsychological battery (cNPB) assessment (See Supplementary methods). Patients with cognitive alterations were re-evaluated at 15 days, 6 months, and 12 months post-stroke using both the cNPB and screening battery. Patients with affective alterations (depression and/or apathy) were followed up at 6- and 12-months post-stroke by the same screening battery. Patients without alterations at baseline were re-evaluated via phone assessment at 1-year post-stroke.

On the same day as the screening, a blood sample was collected through standard venipuncture. Biochemical profiles were obtained from serum samples in mg/dL. Subsequently, *APOE* genotyping was performed using buffy coat (Detailed sample processing is provided in Supplementary methods).

### Biomarker discovery

#### Neuroimaging

MRI was performed within 1 week of hospital admission (5,0 [SD: 1,9] days). Patients with an early recurrence before MRI were excluded. A senior neuroradiologist (RM), who was blind to the clinical features, established the presence and location of any diffusion weighted imaging abnormalities. Ischemic lesions were defined as single or multiple lesions, along with the affected territory and their location. The presence of hemorrhagic transformation, microbleeds, the number of chronic infarcts and, leukoaraiosis were also recorded. IntelliSpace Portal imaging software (version 10.1) was used to calculate the total volume of the hyperintense acute injury in diffusion sequence (cm^3^ units).

#### Metabolomics and lipidomics approach

The experimental design consisted of two phases: discovery and validation. In the discovery phase, plasma samples collected at baseline from MS patients were used (n=25): patients without alterations (n=13), patients with affective alterations (n=6) and patients with cognitive alterations (n=6). The differentially expressed molecules between the three groups were identified using a non-targeted metabolomics/lipidomics approach. Subsequently, a greater number of plasma samples from the same cohort were analysed in the validation phase. The samples included 65 patients without alterations, 14 patients with affective alterations and 11 patients with cognitive alterations.

#### Sample processing

Non-targeted metabolomic and lipidomic profiling was performed on an Agilent 1290 LC system coupled to an electrospray-ionization quadruple time of flight mass spectrometer (Q-TOF 6520 instruments, Agilent Technologies, Barcelona, Spain). Identities were confirmed based on exact mass, retention time (RT), isotopic distribution and MS/MS spectrum using public databases such as Metlin [15], HMDB [16] and LipidMatch [17]. Metaboanalyst platform [18] was used to perform multivariate statistics (partial least squares discriminant analyses, PLS-DA) of the extracted features (See Supplementary methods).

#### Statistical analysis

Qualitative variables were summarized using frequencies and percentages. Quantitative variables were described as mean and standard deviations (SD) for normally distributed data, and median and interquartile range (IQR) for non-normally distributed data. Normally distributed variables were analyzed using t-test or ANOVA. The Mann-Whitney U-test or Kruskal Wallis’s test was used for non-normally distributed variables. Comparisons were performed by means of the Pearson’s chi-squared test for categorical variables. Once the univariate analysis was performed to detect variables associated with the presence of affective and/or cognitive alterations after MS, multivariate models were adjusted to identify predictors of these alterations. The selection of explanatory variables was carried out one by one forward based on the likelihood ratio test (p-value <0.05). Potential first-order interactions were also explored. For all outcome variables, different models obtained from different candidate variables were explored: only clinical variables (Model A), clinical and biochemical variables (Model B), radiological variables (Model C), clinical, biochemical, and radiological variables (Model D), and clinical, biochemical, radiological and omic variables (Model E). To discriminated between the different groups, the odds ratio (OR) was calculated.

Statistical analyses were performed using GraphPad Prism 6 (GraphPad Software, Inc., La Jolla, CA, USA) and SPSS version 20.0 software (SPSS, Chicago, IL). Statistical significance was considered when p-value <0.05.

## RESULTS

### Cohort characteristics

During recruitment period, a total of 727 ischemic stroke patients were admitted to the HUAV, out of which 178 patients (24.5%) met the criteria for MS. Thirty patients declined to participate and, 30 patients were excluded because they did not meet all the study criteria. Finally, 118 patients (66.3%) were included, and 100 patients (84.7%) completed the 12-months follow-up (Figure 1).

**Figure 1:**
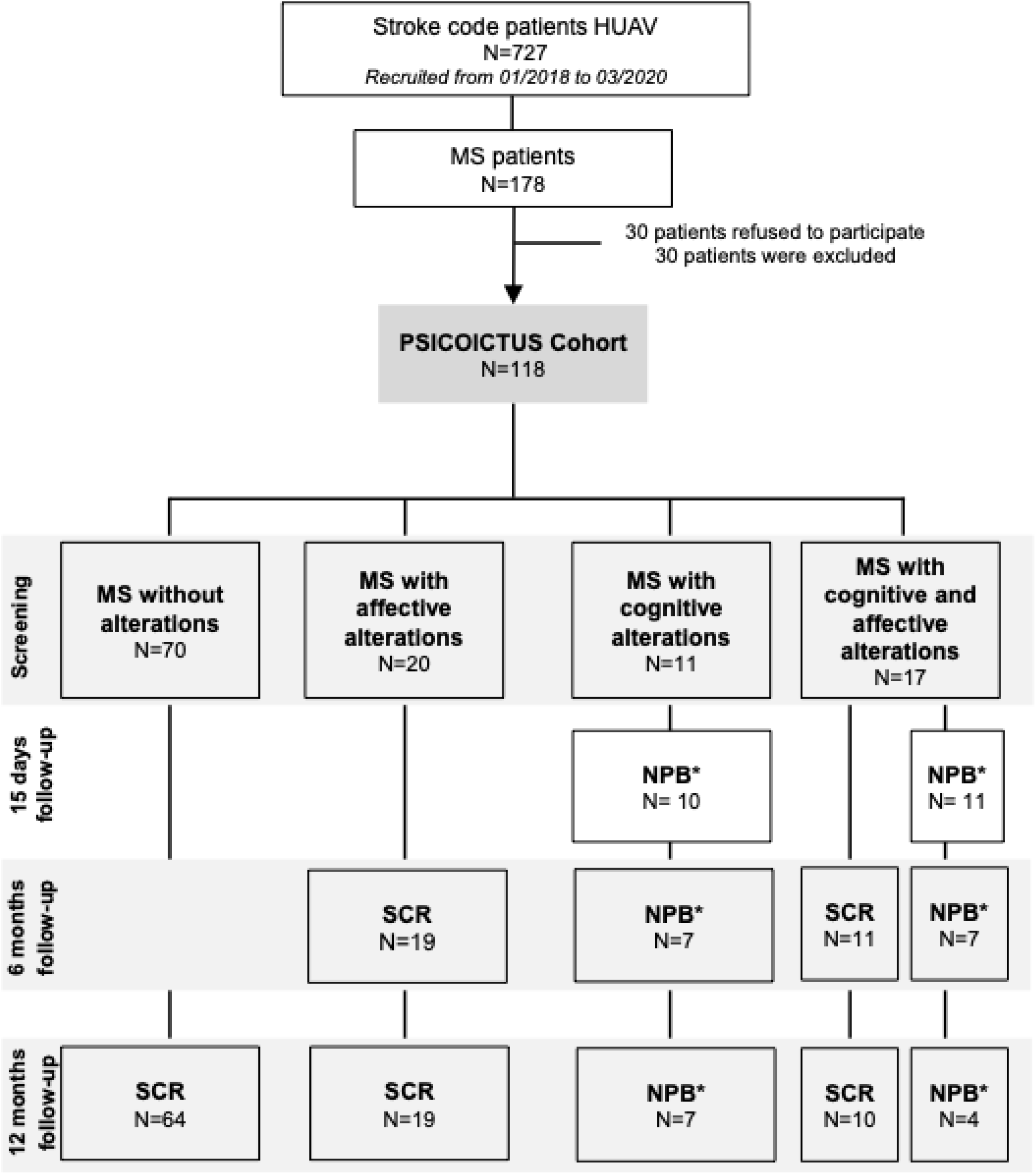
PSICOICTUS cohort flow-chart. Abbreviations: HUAV: Hospital Universitari Arnau de Vilanova; MS: Minor Stroke; SCR: screening; NPB: neuropsychological battery; *: covid time.

The results of the clinical and sociodemographic variables are presented in Table 1. The average age of the cohort was 67.1 years (SD:11.9). MS patients with cognitive alterations were found to be older, while MS patients with affective alterations were younger (75.0 [SD:7.0] and 63.5 [SD:12.2], respectively; *P=0.002*). The study included a higher proportion of men compared to women (81 [68.6%] versus 37 [31.4%]), with men showing a higher prevalence of cognitive alterations and women showing a higher prevalence of affective alterations (*P=0.002*).

**Table 1.** Clinical and sociodemographic variables of the PSICOICTUS cohort according to affective and cognitive profile (baseline data).

|  | All | MS without alterations | MS with affective alterations | MS with cognitive alterations | MS with affective and cognitive alterations | p-value* |
| --- | --- | --- | --- | --- | --- | --- |
| **Total, n** | 118 | 70 | 20 | 11 | 17 |  |
| **Age, mean (SD)** | 67.1(11.9) | 65.2(12.0) | 63.5(12.2) | 75(7.0) | 73.8(9.3) | **0.002** |
| **Male, n (%)** | 81(68.6) | 57(81.4) | 10(50) | 7(63.6) | 7(41.2) | **0.002** |
| **Civil status, n (%)** |  |  |  |  |  |  |
| Married/couple | 87(73.3) | 50(71.4) | 14(70) | 10(90.9) | 13(76.5) | 0.202 |
| Widowed/separated | 31(27.3) | 20(28.6) | 6(30) | 1(9.1) | 4(23.5) |  |
| **Work situation, n (%)** |  |  |  |  |  |  |
| Active | 30(25.4) | 25(35.7) | 5(25) | 0(0) | 0(0) | **0.002** |
| Unemployment | 4(3.4) | 2(2.8) | 1(5) | 2(18.2) | 2(11.8) |  |
| Inability | 6(5.1) | 2(2.9) | 4(20) | 0(0) | 0(0) |  |
| Retired | 78(66.1) | 41(58.6) | 10(50) | 9(81.8) | 15(88.2) |  |
| **Education, n (%)** |  |  |  |  |  |  |
| Primary ed. not completed | 23(19.5) | 10(14.3) | 4(20) | 4(36.4) | 5(29.4) | 0.245 |
| Primary/Secondary education | 81(68.6) | 49(70) | 13(65) | 7(63.6) | 5(29.4) |  |
| Higher education | 14(11.9) | 11(15.7) | 3(15) | 0(0) | 0(0) |  |
| **Live together, n (%)** |  |  |  |  |  |  |
| Partner and/or children | 85(72) | 50(71.4) | 12(60) | 10(90.9) | 13(76.5) | 0.281 |
| Another relative | 12(10.2) | 5(7.1) | 3(15) | 1(9.1) | 3(17.6) |  |
| Alone | 21(17.8) | 15(21.4) | 5(25) | 0(0) | 1(5.9) |  |
| **Caregiver, n (%)** |  |  |  |  |  |  |
| Partner | 73(61.9) | 44(62.9) | 14(70) | 8(72.7) | 7(41.2) | 0.056 |
| Another relative | 29(24.6) | 15(21.4) | 2(10) | 3(27.3) | 9(52.9) |  |
| None | 16(13.6) | 11(15.7) | 4(20) | 0(0) | 1(5.9) |  |
| **Family history, n (%)** |  |  |  |  |  |  |
| Neurological | 28(23.7) | 15(21.4) | 5(25) | 4(36.4) | 4(23.5) | 0.755 |
| Vascular risk | 56(47.5) | 34(48.6) | 12(60) | 2(18.2) | 8(47.1) | 0.166 |
| **Occasional alcohol, n (%)** | 55(46.6) | 40(57.1) | 7(35) | 3(27.3) | 5(29.4) | **0.049** |
| **Smoking, n (%)** |  |  |  |  |  |  |
| Active smoker | 27(22.9) | 20(28.6) | 3(15) | 2(18.2) | 2(11.8) | 0.199 |
| Ex-smoker | 40(33.9) | 25(35.7) | 7(35) | 5(45.5) | 3(17.6) |  |
| **Hypertension, n (%)** | 98(83.1) | 56(80) | 16(80) | 10(90.9) | 16(94.1) | 0.465 |
| **Diabetes mellitus, n (%)** | 42(35.6) | 26(37.1) | 7(35) | 1(9.1) | 8(47.1) | 0.219 |
| **Hyperlipidemia, n (%)** | 72(61) | 45(64.3) | 11(55) | 3(27.3) | 13(76.5) | 0.222 |
| **Atrial fibrillation, n (%)** | 12(10.2) | 5(7.1) | 3(15) | 3(27.3) | 1(5.9) | 0.166 |
| **Previous stroke, n (%)** | 12(10.2) | 7(10) | 2(10) | 1(9.1) | 2(11.8) | 0.996 |
| **Anxiety before stroke, n (%)** | 25(21.2) | 11(15.7) | 9(45) | 1(9.1) | 4(23.5) | **0.028** |
| **Depression before stroke, n (%)** | 24(20.3) | 7(10) | 9(45) | 0(0) | 8(47.1) | **<0.001** |
| **Previous psychiatric treatment, n (%)** | 13(11) | 2(2.9) | 6(30) | 0(0) | 5(29.4) | **<0.001** |
| **APOE genotype** |  |  |  |  |  |  |
| 2/2 | 1(0.8) | 0(0) | 1(5) | 0(0) | 0(0) | 0.386 |
| 2/3 | 15(12.7) | 10(14.3) | 1(5) | 2(18.2) | 2(11.8) |  |
| 3/3 | 76(64.4) | 43(61.4) | 16(80) | 8(72.7) | 9(52.9) |  |
| 3/4 | 17(14.4) | 12(17.1) | 1(5) | 1(9.1) | 3(17.6) |  |
| 4/4 | 0(0) | 0(0) | 0(0) | 0(0) | 0(0) |  |
| Missing data | 9(7.6) | 5(7.1) | 1(5) | 0(0) | 3(17.6) |  |
| **Biochemical profile, mean (SD)** |  |  |  |  |  |  |
| Creatinine | 0.9(0.5) | 0.9(0.3) | 0.8(0.2) | 1.4(1.4) | 0.8(0.3) | **0.009** |
| Triglyceride | 140(53.1) | 151(51.7) | 127(51.3) | 106(25.8) | 132(63.8) | **0.029** |
| **Stroke severity, IQR** |  |  |  |  |  |  |
| **NIHSS** |  |  |  |  |  |  |
| At screening | 2.0<br>[1.0-3.0] | 2.0<br>[1.0-3.0] | 2.0<br>[1.0-3.0] | 1.0<br>[1.0-3.0] | 2.0<br>[1.0-3.0] | 0.717 |
| **mRS** |  |  |  |  |  |  |
| At screening | 1.0<br>[1.0-2.0] | 1.0<br>[1.0-2.0] | 2.0<br>[1.0-2.0] | 1.0<br>[1.0-2.0] | 2.0<br>[1.0-2.0] | 0.348 |
| **Stroke etiologies (TOAST), n (%)** |  |  |  |  |  |  |
| LAA | 23(19.5) | 14(20) | 5(25) | 3(27.3) | 1(5.9) | 0.382 |
| Cardioembolic | 24(20.3) | 11(15.7) | 4(20) | 5(45.5) | 4(23.5) |  |
| SV | 19(16.1) | 13(18.6) | 2(10) | 1(9.1) | 3(17.6) |  |
| Undetermined | 52(44.1) | 32(45.7) | 9(45) | 2 (18.2) | 9(52.9) |  |
| **Stroke treatment, n (%)** |  |  |  |  |  |  |
| Thrombectomy | 0(0) | 0(0) | 0(0) | 0(0) | 0(0) | 0.578 |
| Fibrinolysis | 10(8.5) | 5(7.1) | 2(10) | 1(0.9) | 2(11.8) |  |
| Both | 2(1.7) | 1(1.4) | 0(0) | 1(0.9) | 0(0) |  |
| None | 106 (89.8) | 64 (91.4) | 18 (90) | 9 (81.8) | 15 (88.23) |  |

Abbreviations: MS: minor stroke; SD: standard deviation; ed: education; APOE: apolipoprotein E; IQR: interquartile range; NIHSS: National institute of Health Stroke Scale; mRS: modified Rankin Scale; TOAST: Trial of ORG-10172 in Acute Stroke Treatment; LAA: large artery atherosclerosis; SV: small vessel. \*: p-value

Hypertension was identified as the main risk factor in 98 (83.1%) of the cases. Patients who consumed alcohol occasionally (*P=0.049*), had a history of anxiety (*P=0.028*) or depression (*P≤0.001*) prior to the stroke, and had previously received psychiatric treatment (*P=0.002*) were more likely to experience affective alterations after MS. The group with both affective and cognitive alterations had a higher proportion of retired patients compared to the other groups (n=15 [88.2%]; *P=0.002*).

Regarding the biochemical profile, MS patients with cognitive alterations exhibited higher creatinine concentration (1.4 mg/dL, *P=0.009*) and lower triglyceride concentration (106.0 mg/dL, *P=0.029*).

No significant differences were observed between the groups in terms of APOE genotyping, stroke severity (NIHSS), functionality after stroke (mRS), or the treatment received during the acute phase. Only 10.2% (n=12) of the patients received reperfusion treatment.

### Description and evolution of depression and apathy post-minor stroke

At baseline, 20 patients (17.0%) exhibited symptoms of depression and/or apathy, while 17 patients (14.4%) experienced both affective and cognitive alterations (Figure 1). The affective alterations group had median MADRS and AES-C scores of 11.0 (IQR=8.0-17.0) and 36.5 (IQR=30.0-45.0), respectively. The group with affective and cognitive alterations, the median MADRS and AES-C scores were 11.0 (IQR=7.0-15.0) and 45.0 (IQR=38.0-47.0) (Figure 2A, 2B). At the 12-month follow-up, both groups showed a functional improvement of the affective profile.

**Figure 2:**
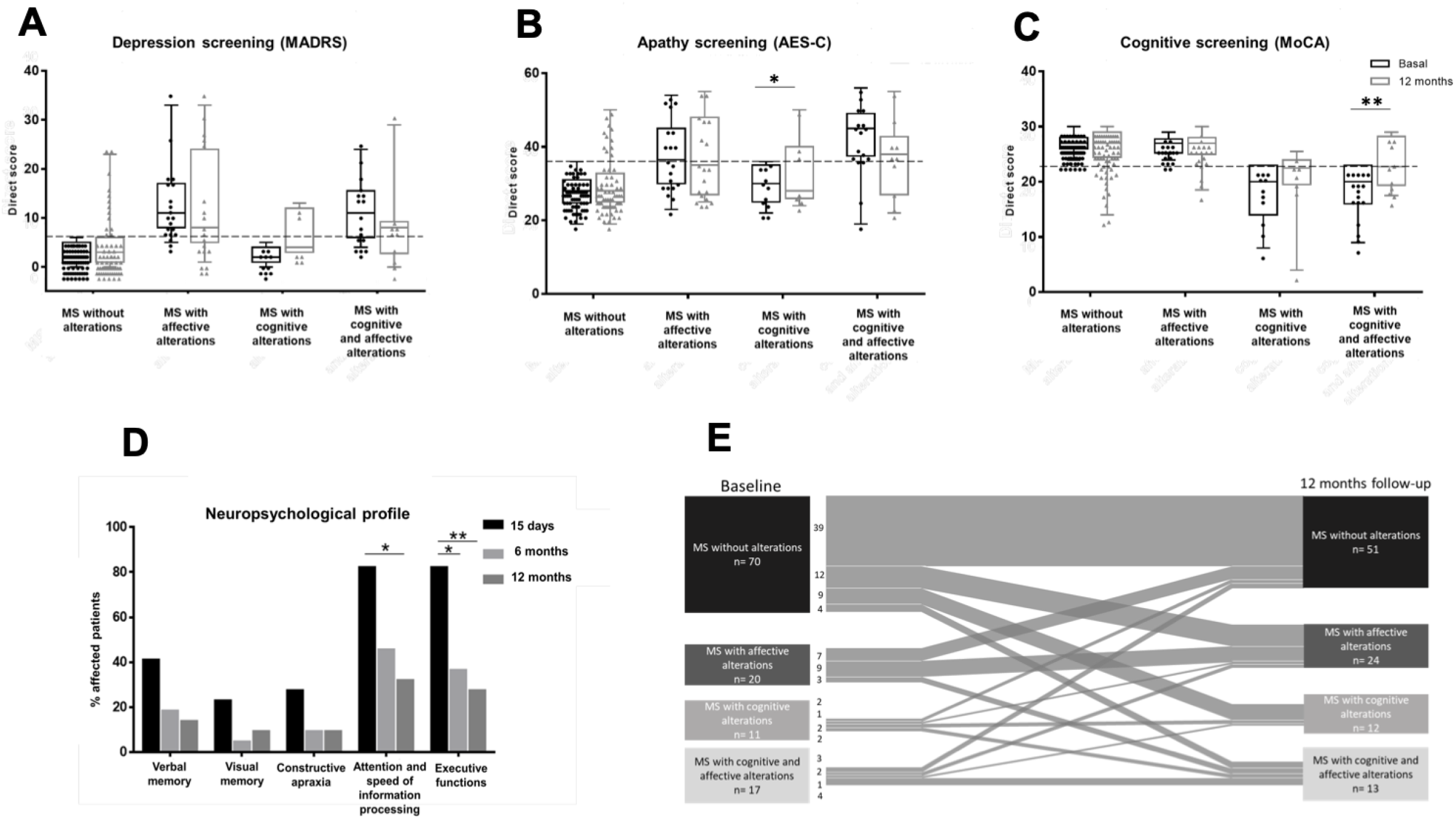
Screening test scores at baseline and 12 months post-stroke, neuropsychological profile. (A) Evolution of post-MS depression according to MADRS; (B) Evolution of post-MS apathy according to AES-C; (C) Evolution of post-MS cognitive alterations according to MoCA; (D) Neuropsychological profile at 15 days, 6 months, and 12 months post-MS; (E) Migration flow of the patients included in the PSICOICTUS cohort. Abbreviations: MS: Minor Stroke; MADRS: Montgomery-Asberg Depression rating scale; AES-C: apathy evaluation scale-clinician; MoCA: Montreal Cognitive assessment; *p-value<0.05; **p-value<0.01.

### Description and evolution of cognitive decline post-minor stroke

At baseline, 11 patients (9.3%) exhibited cognitive alterations, while 17 patients (14.4%) showed both affective and cognitive alterations. The median score on the MoCA test was 20.0 (IQR=16.0-22.5) for the group with cognitive alterations and 21.0 (IQR=18.0-23.0) for the group with affective and cognitive alterations (Figure 2.C). Notably, at the 12-month follow-up, patients with both alterations (n=11) demonstrated significantly improved MoCA scores compared to their baseline: 21.0 (IQR=16.0-23.0) vs. 22.8 (IQR=19.4-28.3; *P=0.028*). Among patients with cognitive decline at baseline, a comprehensive neuropsychological battery was also administered. The most affected cognitive domains were attention and information processing speed (90.4%) and executive functions (90.4%). Over time, there was a progressive improvement across all cognitive domains, with significant improvements observed in the two most affected domains (Figure 2D).

Figure 2E illustrates the distribution of patients based on affective and cognitive screening at baseline and at 12-month follow-up. At the 12-month mark post-MS, 39.1% (n=25) of patients without initial alterations exhibited worsened affectation, while 33.3% (n=12) of patients showed improvement. Furthermore, 41.7% of patients maintained their baseline alterations at 12-month. Ultimately, 49.0% (n=49) of the patients presented some form of alteration at the 12-months follow-up, with affective alterations being the most prevalent (24.0%).

### Radiological profile of affective and cognitive alterations after minor stroke

MRI data were obtained from 93 patients (78.8%) of the cohort. The group with cognitive alterations following MS exhibited the largest infarct volume, followed by the group both affective and cognitive alterations (10.9 [IQR=2.8-23.6] and 8.1 [1.4-9.0]; *P=0,025*) (Figure 3A). Among patients with cognitive alterations post-MS, the majority had acute ischemic lesions affecting the cortical middle cerebral artery (MCA) territory (75.0%) (Figure 3B), and a higher incidence of hemorrhagic transformation (62.5%; *P=0.005*). The group with affective and cognitive alterations showed a higher number of chronic stroke (3.7 [SD:4.7]) and a tendency to have more microbleeds (63.7%; *P=0.069*, Table 2).

**Figure 3:**
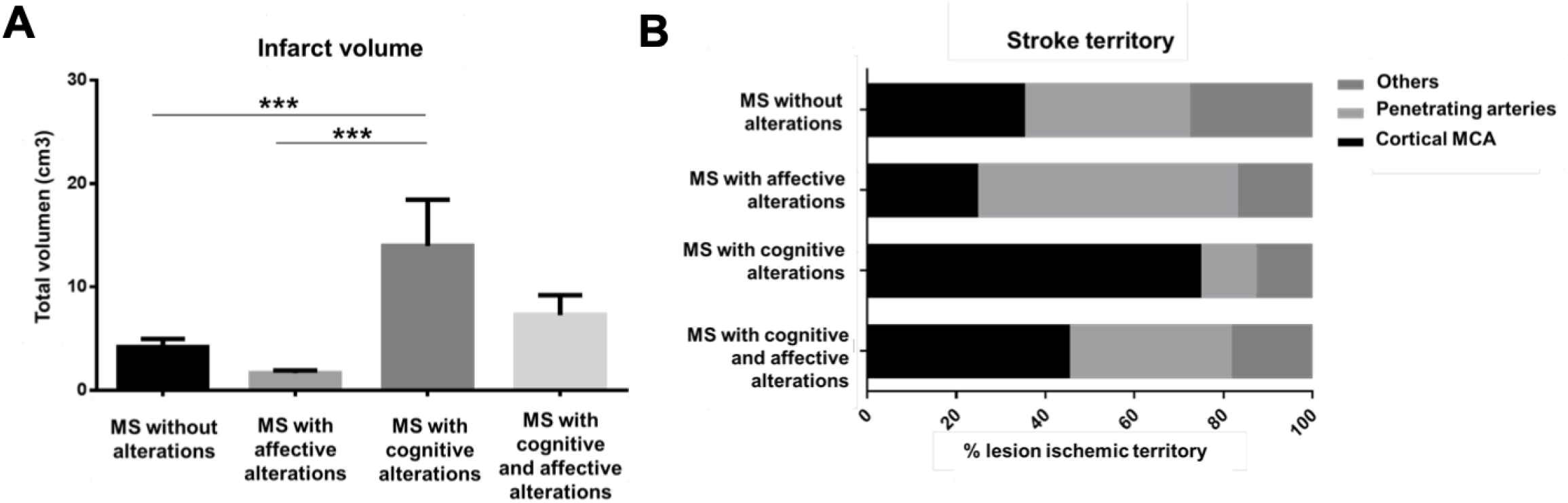
Neuroimaging profile. (A) Differences in infarct volume (cm3) and (B) % territory injury between different groups. Abbreviations: MS: minor stroke; MCA: middle cerebral artery; ***p-value<0.001.

**Table 2.**
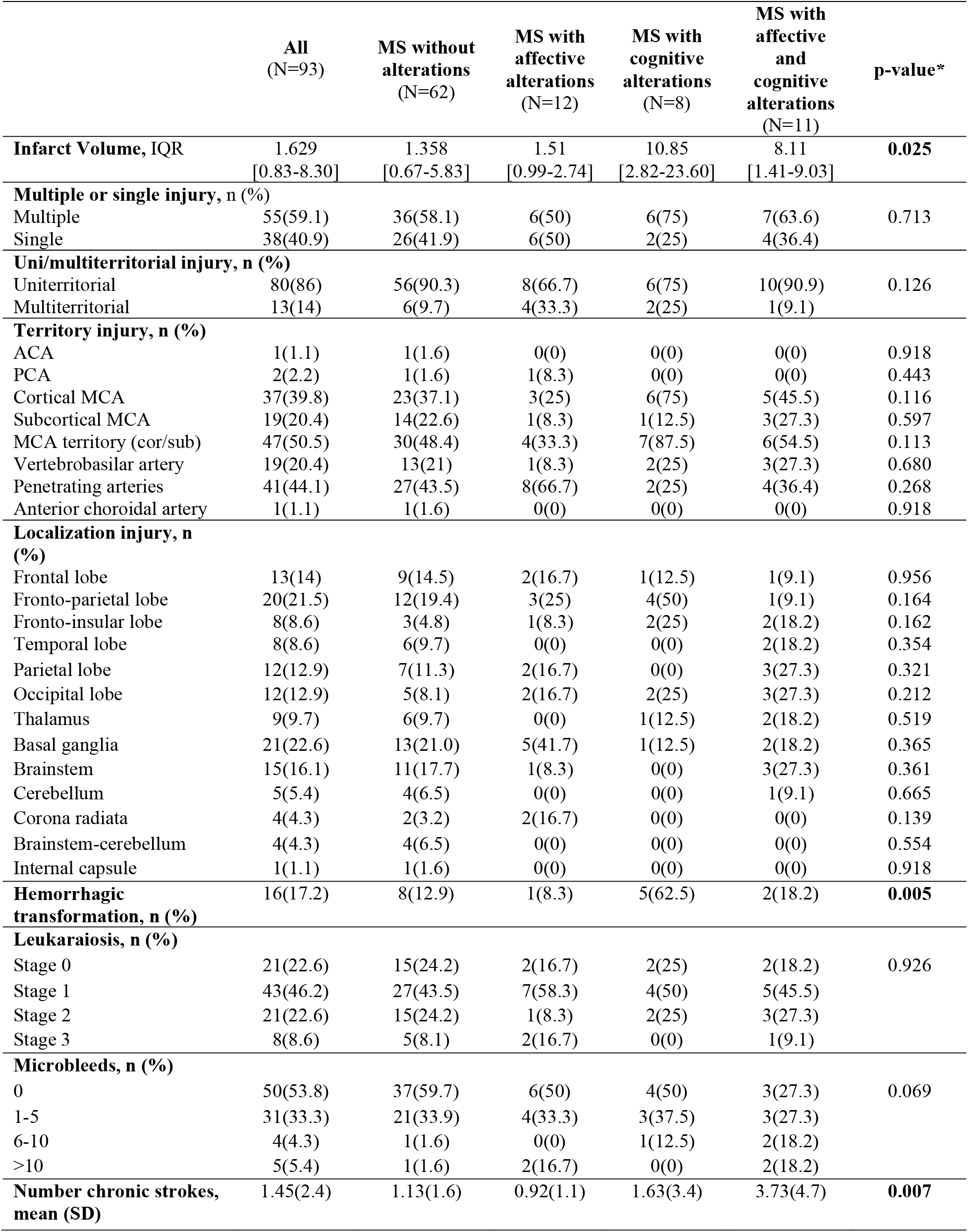
Neuroimaging signature characteristics of the PSICOICTUS cohort according to affective and cognitive profile.

|  | All<br>(N=93) | MS without<br>alterations<br>(N=62) | MS with<br>affective<br>alterations<br>(N=12) | MS with<br>cognitive<br>alterations<br>(N=8) | MS with<br>affective<br>and<br>cognitive<br>alterations<br>(N=11) | p-value* |
| --- | --- | --- | --- | --- | --- | --- |
| **Infarct Volume, IQR** | 1.629<br>[0.83-8.30] | 1.358<br>[0.67-5.83] | 1.51<br>[0.99-2.74] | 10.85<br>[2.82-23.60] | 8.11<br>[1.41-9.03] | **0.025** |
| **Multiple or single injury, n (%)** |  |  |  |  |  |  |
| Multiple | 55(59.1) | 36(58.1) | 6(50) | 6(75) | 7(63.6) | 0.713 |
| Single | 38(40.9) | 26(41.9) | 6(50) | 2(25) | 4(36.4) |  |
| **Uni/multiterritorial injury, n (%)** |  |  |  |  |  |  |
| Uniterritorial | 80(86) | 56(90.3) | 8(66.7) | 6(75) | 10(90.9) | 0.126 |
| Multiterritorial | 13(14) | 6(9.7) | 4(33.3) | 2(25) | 1(9.1) |  |
| **Territory injury, n (%)** |  |  |  |  |  |  |
| ACA | 1(1.1) | 1(1.6) | 0(0) | 0(0) | 0(0) | 0.918 |
| PCA | 2(2.2) | 1(1.6) | 1(8.3) | 0(0) | 0(0) | 0.443 |
| Cortical MCA | 37(39.8) | 23(37.1) | 3(25) | 6(75) | 5(45.5) | 0.116 |
| Subcortical MCA | 19(20.4) | 14(22.6) | 1(8.3) | 1(12.5) | 3(27.3) | 0.597 |
| MCA territory (cor/sub) | 47(50.5) | 30(48.4) | 4(33.3) | 7(87.5) | 6(54.5) | 0.113 |
| Vertebrobasilar artery | 19(20.4) | 13(21) | 1(8.3) | 2(25) | 3(27.3) | 0.680 |
| Penetrating arteries | 41(44.1) | 27(43.5) | 8(66.7) | 2(25) | 4(36.4) | 0.268 |
| Anterior choroidal artery | 1(1.1) | 1(1.6) | 0(0) | 0(0) | 0(0) | 0.918 |
| **Localization injury, n (%)** |  |  |  |  |  |  |
| Frontal lobe | 13(14) | 9(14.5) | 2(16.7) | 1(12.5) | 1(9.1) | 0.956 |
| Fronto-parietal lobe | 20(21.5) | 12(19.4) | 3(25) | 4(50) | 1(9.1) | 0.164 |
| Fronto-insular lobe | 8(8.6) | 3(4.8) | 1(8.3) | 2(25) | 2(18.2) | 0.162 |
| Temporal lobe | 8(8.6) | 6(9.7) | 0(0) | 0(0) | 2(18.2) | 0.354 |
| Parietal lobe | 12(12.9) | 7(11.3) | 2(16.7) | 0(0) | 3(27.3) | 0.321 |
| Occipital lobe | 12(12.9) | 5(8.1) | 2(16.7) | 2(25) | 3(27.3) | 0.212 |
| Thalamus | 9(9.7) | 6(9.7) | 0(0) | 1(12.5) | 2(18.2) | 0.519 |
| Basal ganglia | 21(22.6) | 13(21.0) | 5(41.7) | 1(12.5) | 2(18.2) | 0.365 |
| Brainstem | 15(16.1) | 11(17.7) | 1(8.3) | 0(0) | 3(27.3) | 0.361 |
| Cerebellum | 5(5.4) | 4(6.5) | 0(0) | 0(0) | 1(9.1) | 0.665 |
| Corona radiata | 4(4.3) | 2(3.2) | 2(16.7) | 0(0) | 0(0) | 0.139 |
| Brainstem-cerebellum | 4(4.3) | 4(6.5) | 0(0) | 0(0) | 0(0) | 0.554 |
| Internal capsule | 1(1.1) | 1(1.6) | 0(0) | 0(0) | 0(0) | 0.918 |
| **Hemorrhagic transformation, n (%)** | 16(17.2) | 8(12.9) | 1(8.3) | 5(62.5) | 2(18.2) | **0.005** |
| **Leukaraiosis, n (%)** |  |  |  |  |  |  |
| Stage 0 | 21(22.6) | 15(24.2) | 2(16.7) | 2(25) | 2(18.2) | 0.926 |
| Stage 1 | 43(46.2) | 27(43.5) | 7(58.3) | 4(50) | 5(45.5) |  |
| Stage 2 | 21(22.6) | 15(24.2) | 1(8.3) | 2(25) | 3(27.3) |  |
| Stage 3 | 8(8.6) | 5(8.1) | 2(16.7) | 0(0) | 1(9.1) |  |
| **Microbleeds, n (%)** |  |  |  |  |  |  |
| 0 | 50(53.8) | 37(59.7) | 6(50) | 4(50) | 3(27.3) | 0.069 |
| 1-5 | 31(33.3) | 21(33.9) | 4(33.3) | 3(37.5) | 3(27.3) |  |
| 6-10 | 4(4.3) | 1(1.6) | 0(0) | 1(12.5) | 2(18.2) |  |
| >10 | 5(5.4) | 1(1.6) | 2(16.7) | 0(0) | 2(18.2) |  |
| **Number chronic strokes, mean (SD)** | 1.45(2.4) | 1.13(1.6) | 0.92(1.1) | 1.63(3.4) | 3.73(4.7) | **0.007** |

Abbreviations: MS: minor stroke; IQR: interquartile range; Uni: uniterritorial; ACA: Anterior cerebral artery; PCA: Posterior cerebral artery; MCA: Middle cerebral artery; cor: cortical; sub: subcortical. \*: p-value

### Circulating biomarkers of affective and cognitive alterations after minor stroke

A specific metabolic (Figure 4A) and lipidic (Figure 4B) was identified with good precision for each experimental group included in the discovery phase. By an univariate statistical analysis, it was observed that there were 6 metabolites (2-Hydroxyhexadecanoylcarnitine, DG (36:7), PE (13D5/13M5), PE-NMe (40:4), Tocophersolan, and PG (34:0)), and 1 lipid; (PC-P (38:4)) that exhibited differential expression between MS patients without alterations and MS patients with cognitive alterations. Furthermore, 6 metabolites (DG (36:7), PS (40:6), Isoleucyl-Isoleucine, PC (38:4), PG (34:0), and N-Decanoylglycine) and 2 lipids (PC-P (38:4), and PC (38:2)) were found to be differentially expressed between MS patients without alterations and those with affective alterations (Figure 4C).

**Figure 4:**
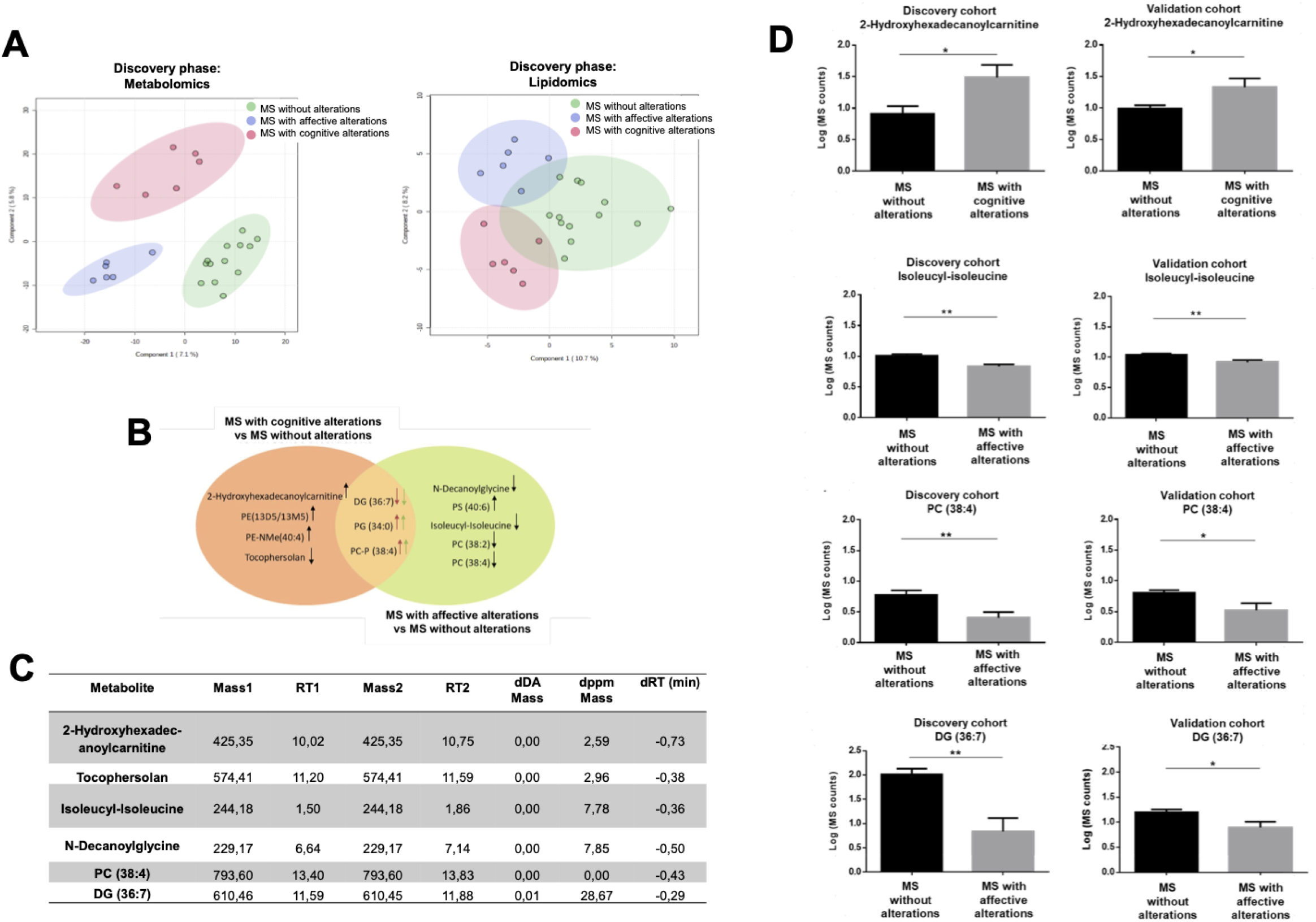
Circulating biomarker profile. (A) PLS-DA graph showing the profile of plasma metabolites/lipids according to the affective and cognitive function profile of the patients included in the discovery phase. (B) Metabolites and lipids differentially expressed in the discovery phase in MS with cognitive alterations group and in MS with affective alterations group compared to the MS without alterations group. The arrows indicate whether they are up-or down-regulated. (C) Comparison of the masses and RTs of the identified metabolites obtained in the discovery phase (Mass1/RT1) and the validation phase (Mass2/RT2). (D) Quantification of the expression of the metabolites identified in the discovery phase and the validation phase that are differentially expressed between groups. Abbreviations: MS: Minor Stroke; PE: phosphatidylethanolamine; PS: phosphatidylserine; PC: phosphatidylcholine; DG: diacylglyceride; PG: phosphoglyceride; RT: retention time; d: difference; DA: Daltons; ppm: parts per million; *p-value<0.05; **p-value<0.01.

Subsequently, a validation phase was conducted, showing the mass values and RTs of the identified metabolites from both discovery and validation phases (Figure 4D). Among the metabolites and lipids identified and differentially expressed in the discovery phase, 2-Hydroxyhexadecanoylcarnitine was confirmed as upregulated in patients with MS and cognitive alterations compared to the MS without alterations group. On the other hand, Isoleucyl-Isoleucine, DG (36:7) and PC (38:4) were confirmed as downregulated in patients with MS and affective alterations compared to the MS without alterations group (Figure 4E).

### Clinical, biochemical, radiological and omic signatures predictors of cognitive and affective alterations post-minor stroke

Factors associated with cognitive alterations after MS were age older 70 years (OR:15.9; CI95% 1.9-131.5; *P=0.010* [Model A], OR:14.6; CI95% 1.6-133.8; *P=0.023* [Model B], OR:11.5; CI95% 1.2-112.9; *P=0.036* [Model D] and OR:34.3; CI95% 2.2-545.6; *P=0.012* [Model E]), higher creatinine levels (OR:2.6; CI95% 0.9-8.1; *P=0.090* [Model D]), MCA cortical territory injury (OR:5.5; CI95% 1.0-29.4; *P=0,048* [Model C], OR:11.5; CI95% 1.4-98.4; *P=0.025* [Model D] and OR:17.1; CI95% 1.5-191.3; *P=0.021* [Model E]) and concentrations greater than 1.05 of 2-Hydroxyhexadecanoylcarnitine (OR:11.0; CI95% 1.1-113.8; *P=0.044* [Model E]). Higher triglyceride levels behaved as a protective factor (OR:0.9; CI95% 0.9-1.0; *P=0.023* [Model B]).

Factors associated with affective alterations after MS were female sex (OR:3.3; CI95% 1.0-10.7; *P=0.043* [Model A] and OR:4.5; CI95% 1.1-18.6; *P=0.038* [Model D]), medical record of depressive syndrome (OR:7.4; CI95% 1.1-25.1; *P=0.001* [Model A], OR:10.0; CI95% 2.9-34.8; *P<0.001* [Model B], OR:6.6; CI95% 1.5-29.3; *P=0.013* [Model D] and OR:6.9; CI95% 1.8-26.4; *P=0.005* [Model E]), concentrations less than 0.90 of Isoleucyl-Isoleucine (OR:5.5; CI95% 1.5-20.2; *P=0.011* [Model E]) and concentrations less than 1.19 of PC (38:4) (OR:4.5; CI95% 1.2-16.6; *P=0.023* [Model E]).

Finally, factors associated with affective and cognitive alterations after MS were female sex (OR:6.3; CI95% 1.8-21.9; *P=0.004* [Model A], OR:7.1; CI95% 2.5-44.4; *P=0.004* [Model B] and OR:15.8; CI95% 2.5-100.3; *P=0.001* [Model D]), medical record of depressive syndrome (OR:8.0; CI95% 2.1-31.0; *P=0.003* [Model A], OR:10.5; CI95% 2.5-44.4; *P=0.001* [Model B] and OR:6.4; CI95% 0.9-43.9; *P=0.058* [Model D]) and higher number of chronic stroke (OR:1.5; CI95% 1.1-2.1; *P=0.015* [Model C] and OR:1.5; CI95% 1.0-2.3; *P=0.034* [Model D]).

## DISCUSSION

Despite affective and cognitive impairment are crucial factors that impact the functional recovery of stroke patients, only a few studies have specifically investigated the predictors of affective and cognitive alterations in patients with MS. Our study identified several factors associated with cognitive and affective alterations in patients with MS. These findings emphasize the impact of age, specific biomarkers, and clinical characteristics on the development of these alterations. Although depression and apathy are common sequelae of stroke, there is uncertainty regarding their incidence, relevance, and correlation with the location of the lesion in MS. In our study, baseline assessment revealed 31.4% of patients exhibited affective alterations, either alone (17.0%) or in conjunction with cognitive decline (14.4%). During the one-year follow-up, it was observed that the group of MS patients with affective alterations, as well as the group with affective and cognitive alterations, showed a tendency towards improvement. Longitudinal studies on post-stroke depression have described a dynamic natural progression with improvements over time, but there is still limited information on whether the course of depression post-stroke (DPI) differs from depression without a history of stroke [19]. As for apathy, it remained stable in the group with affective alterations and tended to improve in the group with affective and cognitive alterations. This may be attributed to challenges in accurately diagnosing and treating apathy. On the other hand, cognitive function showed improvement in the MS and affective and cognitive alterations group, possibly explained by the reduction in depression and apathy in these patients [6].

A higher proportion of women was observed among MS patients with affective alterations, and female sex emerged as a predictor for experiencing such alterations in the multivariate analysis. Wade and colleagues were the first authors to observe the association between female sex and DPI [20]. Being female was a significant risk factor for DPI during the acute and subacute phase [21]. Hormonal changes have been proposed as a potential mechanism contributing to the occurrence of DPI, as they can impact on women’s mood [22]. Significant differences were found in previous psychiatric history and psychopharmacological treatment among the different groups, particularly in relation to a history of depressive syndrome, which emerged as a predictive factor for affective alterations in the multivariate analysis. This relationship is supported by the existing literature, as a history of depression or mental disorders were risk factors for DPI [23] [24].

Unfortunately, previously no biochemical, or radiological variables were identified as predictors for experiencing affective alterations after MS. In our study, the analysis of circulating biomarkers showed a relationship between some metabolites and the presence of affective alterations after MS. Specifically, concentrations of Isoleucyl-Isoleucine below 0.9 and concentrations of PC (38:4) below 1.2 emerged as predictors for affective alterations after MS. To the best of our knowledge, there are no previous studies linking these two metabolites to ischemic stroke or the development of affective disturbances such as depression and apathy following a stroke.

In the present study, a total of 11 patients (9.3%) exhibited cognitive alterations, while 17 patients (14.4%) experienced affective and cognitive alterations following MS. Cognitive dysfunction was observed in all subjects who underwent a cNPB at baseline, with attention, processing speed, and executive functions being the most affected domains. A strong association between multi-domain non-amnesic cognitive impairment and vascular pathology had been described [7].

In the longitudinal one-year follow-up, a higher prevalence of depression was observed in MS patients with cognitive alterations. The predictive value of cognitive impairment during the acute post-stroke phase as a long-term risk factor for the development of depressive symptoms and a lower quality of life was described [25]. In the same group of patients, a decrease in apathy, associated with a cognitive improvement, was also observed at 12-months post-MS [26]. This relationship can be explained by the inability to engage in goal-oriented thinking could lead to a loss of interest and lack of effort when performing cognitive tests, resulting in lower scores [27]. Advanced age has emerged as a significant predictor of cognitive impairment following MS. Over the past few decades, there has been a substantial increase in life expectancy, leading to a rise in the prevalence of dementia and cognitive issues among the adult population [28].

In our analysis of the patients’ biochemical profiles, high levels of creatinine were predictive of cognitive alterations following MS. Generally, elevated blood creatinine levels and reduced urinary levels indicate kidney disease [29]. In individuals with advanced chronic kidney disease, nervous system dysfunctions contribute significantly to disability. These cognitive impairments manifest as disruptions in global cognitive function, executive function, language, memory, attention, mental fatigue, and affective disorders [30]. The identification of advanced age as a risk factor for post-MS cognitive impairment, along with the predictive value of high creatinine levels, underscores the importance of considering these factors in the management and treatment of individuals who have experienced MS. Efforts to address cognitive impairments and mitigate their impact on patients’ lives can lead to improved outcomes and enhanced quality of life.

The literature on the relationship between post-stroke cognitive impairment and neuroimaging findings is controversial. Based on our results, the presence of acute ischemic lesions in the MCA cortical territory emerged as a risk predictor for post-MS cognitive impairment in the multivariate model. The location of the ischemic lesion within the MCA territory has been associated with decreased functional independence following stroke [31]. Additionally, several studies have identified it as a predictive factor for post-stroke deterioration, findings that align with our study. In addition, there is a differential radiological signature between the patients with isolated cognitive impairment or patients with MCI associated with affective disturbances. The group with both alterations exhibited a higher number of chronic ischemic strokes and emerged as a predictor in the multivariate analysis. Mandzia and colleagues had previously observed that patients with previous chronic cortical infarcts had cognitive dysfunction in executive functions and processing speed [9]. On the other hand, according to the results of a meta-analysis carried out in 2017, this type of injury was not associated with presented affective alterations [32]. Furthermore, we discovered and validated that 2-Hydroxyhexadecanoylcarnitine levels were significantly elevated in MS patients with cognitive alterations compared to those without alterations. Plasma concentrations exceeding 1.05 became a predictor for cognitive alterations following MS. Although this metabolite has not been directly linked to stroke or cognitive decline, elevated concentrations have been associated with mitochondrial trifunctional protein deficiency [33]. Mitochondrial dysfunction may contribute to the aberrant expression and processing of β-amyloid precursor protein, thus potentially serving as a driving factor in the development of MCI and and Alzheimer’s disease [34].

The main limitation of this study was the sample size, mainly due to the fact that the study recruitment was abruptly cancelled in March 2020 because of the global COVID-19 pandemic. Moreover, a control group (healthy) was not included, which might contribute to define whether affective and/or cognitive alterations would be related with the ischemic event, or there was a reactive phenomenon.

In conclusion, despite presenting mild neurological symptoms, 40.7% of patients with MS developed affective and cognitive alterations at baseline. Only 10.2% of the patients received reperfusion treatment. This finding underscores the importance of treating these patients during the acute phase, particularly in cases that exhibit predictive risk factors. Patients without alterations, as well as those with affective and/or cognitive alterations, demonstrate distinct neuropsychological, radiological, and circulating metabolic biomarkers profiles. These findings offer a new prognostic strategy and support the implementation of a differential management approach for patients with MS.

## Authors contributions

FP, GP, GA conceived the study. FP, GA, CP designed experiments. FP, GM, DVJ, CP cohorts’ recruitment and clinical data. RM, CP analyzed imaging data. FP, GA, CP participated on data interpretation and draft the manuscript. All authors critically revised the final version of the manuscript. All authors approved the final version to be published. FP procured funding.

## Data Availability Statement

Requests for access to the data reported in this paper will be considered by the Lead contact on reasonable basis.

## Acknowledgements

We are grateful to all recruited patients, the members of Clinical Neuroscience group at IRBLleida, especial thanks to Sara Salvany and Cristina García, and personal of Neurology Department at Hospital Universitari Arnau de Vilanova de Lleida for scientific discussions and instrumental help. We also acknowledge the use of the facility of Biobank (B.0000682), Plataforma Biobancos PT17/0015/0027 and lipidomics SCT at IRBLleida. CP was supported by a grant from Contracte Predoctoral en formació de la Universitat de Lleida (FP: PI17 -01725); INVICTUS plus Research Network (Carlos III Health Institute) (FP, CP, and GA: RD16-0019-0017).

